# mBrainGT: Modular Graph Transformer for Brain Disorder Diagnosis

**DOI:** 10.1101/2024.08.30.24312819

**Authors:** Ahsan Shehzad, Shuo Yu, Dongyu Zhang, Shagufta Abid, Feng Xia

## Abstract

Functional brain networks play an essential role in the diagnosis of brain disorders by enabling the identification of abnormal patterns and connections in brain activities. Previous methods often rely on whole brain functional connectivity approaches to construct these networks using Functional Magnetic Resonance Imaging (fMRI) data. However, these approaches introduce noise and overlook localized disruptions within specific brain subnetworks, leading to potential misdiagnoses. To address this challenging issue, we propose mBrainGT, a modular brain graph transformer model that focuses on modular functional connectivity (mFC) to improve the diagnosis of brain disorders. Compared to existing methods, mBrainGT constructs and analyses functional brain subnetworks individually, reflecting the inherent structure of the brain. It captures both local features within each modular network and their interactions through self-attention and cross-attention mechanisms. It also learns global interactions via adaptive fusion. We validate mBrainGT on three benchmark datasets (ADNI, PPMI, and ABIDE). The results demonstrate that mBrainGT outperforms existing methods in diagnostic accuracy, providing more robust and precise representations of the brain network essential for accurate disease detection. Our study highlights the potential of modular connectivity-based graph learning in the refinement of brain disorder diagnostics, offering a more precise and biologically relevant representation of functional brain networks.

## I. Introduction

**T**HE diagnosis of brain disorders using brain network analysis is a rapidly evolving field of research that holds significant promise for improving medical outcomes [1], [2]. fMRI plays a crucial role in the construction of these brain networks by measuring brain activity and identifying connectivity patterns between different regions [3]. The significance of brain network analysis lies in its potential to provide a more comprehensive understanding of brain function and dysfunction enabling the early detection and diagnosis of various brain disorders such as Alzheimer’s disease (AD), Parkinson’s disease (PD), and Autism pectrum disorder (ASD) [4]–[6] By analyzing alterations in brain network connectivity, researchers and clinicians can identify biomarkers and develop targeted interventions that ultimately lead to more effective treatments and improved patient outcomes.

Numerous efforts have been made to analyze brain networks for the diagnosis of brain disorders. Conventional methods for brain network analysis usually involve constructing brain networks using Pearson correlation coefficients computed from fMRI time-series data [3], [7]. These approaches then extract handcrafted topological features, such as node degrees or clustering coefficients, and apply classical machine learning classifiers [8]. Despite their utility, these methods heavily rely on manual feature selection, which can be suboptimal when dealing with high-dimensional, heterogeneous neuroimaging data. More recently, emerging graph learning techniques, particularly variants of graph neural networks (GNNs), have gained traction for the diagnosis of brain disorders [9], [10]. These models automatically learn salient features from graph-structured input, thus passing extensive manual feature engineering [11], [12]. However, GNNs often exhibit limited receptive fields, which can restrict their ability to capture long-range dependencies crucial for identifying diffuse pathological signatures. Graph transformers have recently emerged as a powerful alternative of GNNs [13]. By exploiting selfattention and related mechanisms, they can capture global dependencies across nodes [14], [15]. This capability holds significant promise for brain network analysis, where comprehensive inter-regional interactions play a pivotal role in disease pathology [16], [17].

Despite the advances in brain network analysis, current graph transformer models still struggle with significant limitations that hinder their diagnostic accuracy. Existing approaches fail to adequately capture localized disruptions within specific brain modules, which are essential for identifying disorder-specific biomarkers. This limitation arises primarily from the reliance on whole-brain connectivity techniques (illustrated in Fig. 1) that lack the capacity to model the interactions between distinct subnetworks in the brain [18], [19]. The negative impact of these issues is profound, as it leads to inaccurate brain network representations, resulting in misdiagnoses and delayed treatment for individuals with neurological disorders. This study hypothesises that a more focused analysis on modular functional connectivity (mFC) will enhance diagnostic accuracy by capturing localized disruptions within key brain networks.

**Fig. 1.**
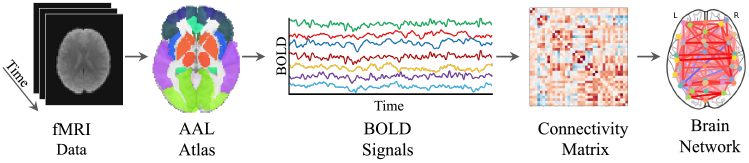
Whole-Brain Connectivity Method: Existing approaches model functional brain networks using whole-brain connectivity. However, they introduce noisy connections and overlook localized disruptions, potentially causing misdiagnosis.

To address these challenges, we propose a modular brain graph transformer model (mBrainGT), that explicitly focuses on mFC to improve brain network analysis for diagnosing brain disorders. In contrast to existing methods, mBrainGT constructs functional brain subnetworks individually, that reflect the brain’s inherent structure. The model then integrates these modular features using self-attention to identify intricate local relationships within each subnetwork and crossattention to understand interactions between different brain subnetworks [20]. An adaptive fusion mechanism further integrates global patterns, enabling the model to capture complex connectivity patterns across the entire brain. We evaluate the performance of mBrainGT through experiments on three benchmark datasets: ADNI, PPMI, and ABIDE. The results demonstrate that mBrainGT surpasses existing methods in diagnostic accuracy, offering more robust and precise brain network representations crucial for accurate disease detection. By providing a holistic global perspective, mBrainGT effectively addresses challenges in whole-brain connectivity, achieving superior performance in brain disease diagnosis and marking a significant advancement in neuroimaging analytics. Our key contributions are summarized as follows:

1. We propose the mBrainGT model to address the limitations of whole-brain connectivity approaches in capturing modular brain connectivity. By focusing on localized disruptions within and between brain subnetworks, mBrainGT reduces noise and enhances diagnostic accuracy for brain disorders.
2. The mBrainGT model is designed to capture functional connectivity within individual brain modules, reflecting the biological organization of brain functions. Using self-attention and cross-attention mechanisms, the model extracts and integrates both intra- and inter-module features, producing a holistic representation that improves the diagnostic capabilities.
3. We validate our model using three benchmark datasets: ADNI, PPMI, and ABIDE, each representing a different brain disease: AD, PD, and ASD, respectively. The experimental results demonstrate that mBrainGT outperforms state-of-the-art methods.

The remainder of this paper is structured as follows: Section II reviews related work and Section III describes the design of mBrainGT, emphasizing its architecture and implementation. Section IV presents and discusses the experimental results, evaluating the performance and implications of the proposed design. Finally, Section V concludes the paper and suggests directions for future research.

## II. Related Work

### A. Brain Network Analysis for Brain Disorder Diagnosis

Brain network analysis has emerged as a powerful tool for understanding and diagnosing brain disorders by capturing patterns of neural connectivity from neuroimaging data [21], [22]. Brain networks, commonly are constructed from fMRI data, model brain regions as nodes and their functional interactions as edges [23], [24]. Typically, the Automated Anatomical Labeling (AAL) atlas [25] defines regions of interest (ROIs), while Pearson correlation coefficients between ROI time-series signals generate connectivity matrices [1], [26]. Brain network analysis methods are broadly classified into graph theory-based and graph learning-based approaches, each with distinct advantages and limitations. Graph theory-based methods, among the earliest approaches, utilise network science metrics such as degree centrality, clustering coefficients, modularity, and small-worldness to detect topological alterations in brain networks [8]. These methods have illuminated disruptions linked to disorders, including Alzheimer’s disease (AD) and autism spectrum disorder (ASD) [27], [28]. However, they rely on handcrafted features, necessitating manual selection of network metrics, which may obscure complex patterns. This manual feature selection introduces subjectivity and reduces generalisability across datasets and disorders [29].

Graph learning-based approaches, particularly GNNs, have emerged to address the limitations of manual feature engineering in brain network analysis [30], [31]. GNNs leverage graph structures to automatically learn node embeddings and connectivity patterns, enhancing brain disorder classification [32], [33]. Li et al. [15] introduced BrainGNN, which incorporates ROI-aware graph convolutional layers and an ROI-selection pooling mechanism to improve interpretability and diagnostic accuracy. Similarly, Cui et al. [9] developed BrainGB, a benchmark framework that standardizes GNN-based method evaluations on brain network datasets, promoting research reproducibility. Luo et al. [34] proposed KDGCN, a knowledge distillation-guided GNN employing a teacher-student framework to enhance prediction accuracy and provide interpretable insights into abnormal brain regions. Despite these advancements, GNN-based models still face a key limitation: their receptive fields are restricted to local neighborhoods due to graph convolution operations. This constraint hampers their ability to capture long-range dependencies within brain networks. Consequently, GNN models struggle to model complex neural interactions spanning multiple functional modules, which are critical for understanding brain disorders.

### B. Graph Transformers

To address the limitations of GNNs, graph transformers have emerged as a promising alternative, leveraging self-attention mechanisms to model both local and global dependencies in graph-structured data [35]. Unlike GNNs, which aggregate information based on node neighborhoods, graph transformers capture interactions between any pair of nodes, making them well-suited for representing complex connectivity patterns within the brain networks [36], [37]. This ability to model long-range dependencies has driven the adoption of graph transformers in various fields, including social networks [38], protein interaction networks [39], and brain disease diagnosis.

Several graph transformer-based models have been applied to brain network analysis, offering new perspectives on neurological disorder diagnosis. Kan et al. [18] introduced BRAINNETTF, a transformer framework employing orthonormal clustering readouts to identify functional brain modules, enhancing interpretability through attention mechanisms. Yu et al. [40] proposed ALTER, integrating a biased random walk strategy with adaptive attention to capture long-range dependencies, achieving improved diagnostic performance. Jun et al. [41] developed the Medical Transformer, designed for volumetric brain imaging, which represents 3D images as 2D slice sequences and employs multi-view spatial encoding with self-supervised pretraining to enhance accuracy and data efficiency. Despite these advances, existing models face limitations due to whole-brain connectivity approaches, which treat the brain as a single interconnected graph. Such methods often overlook modular structures, including the Default Mode, Salience, and Limbic networks, crucial for cognition and behaviour. Ignoring these subnetworks can obscure local disruptions essential for disorder-specific diagnoses. Additionally, whole-brain models introduce noisy connections, reducing representational specificity and diagnostic accuracy. To address these gaps, we propose mBrainGT, a modular brain graph transformer framework explicitly modelling mFC for brain network analysis.

## III. The Design of mBrainGT

### A. Problem Formulation

The goal of this study is to improve the accuracy of brain disorder diagnosis by leveraging mFC representations instead of whole-brain connectivity methods. The computational task is to learn disorder-specific connectivity features from fMRI data, capturing critical disruptions in functional subnetworks and their interactions. Let 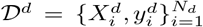 represent the dataset for brain disorder *d*, where 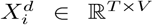 is the fMRI time series for subject *i*, with *T* time points and *V* brain regions, and 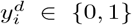 denotes the binary disease label (patient vs. control). The mBrainGT framework aims to learn a function *f*^*d*^ : *X*^*d*^ *→ y*^*d*^ that optimally classifies subjects for a given disorder *d*, where *d* ∈ {AD, PD, ASD}. Given fMRI data 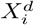, the modular brain network construction module partitions the brain into *M* functional subnetworks using the functional atlas 𝒜_*F*_ (Yeo2011) [42] and anatomical atlas 𝒜_*A*_ (Brainnetome) [43], producing modular brain graphs 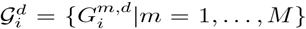, where 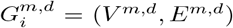. The modular brain graph transformer encodes each 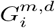 using self-attention within modular encoders and cross-attention in cross-modular encoders to model inter-network interactions. The final hierarchical attention pooling aggregates the representations into a global embedding 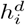, which is mapped to 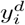 via a classifier 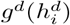. The framework optimizes the classification loss 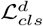 separately for each disorder, ensuring disorder-specific diagnostic accuracy. The framework and workflow of mBrainGT are depicted in Fig. 2.

**Fig. 2.**
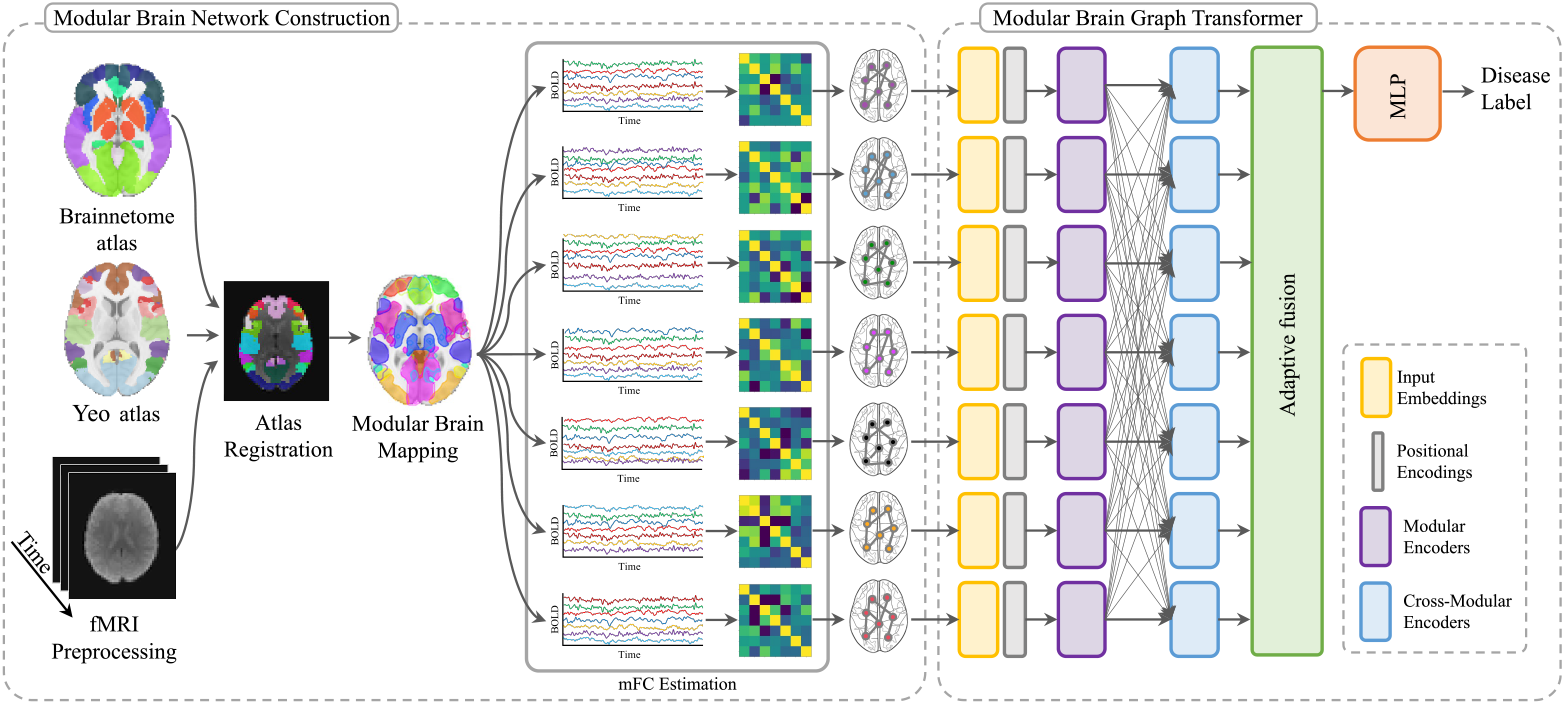
The framework of mBrainGT.

### B. Modular Brain Network Construction

In this study, we construct modular brain networks from raw fMRI data to perform an analysis of brain networks from the perspective of mFC of brain.

#### 1) fMRI Preprocessing

We pre-processed raw fMRI data to eliminate motion artifacts, standardize spatial alignment, and reduced scanner-induced variability, ensuring reliable functional connectivity estimates. The pipeline, implemented using the Nipype framework, integrates interfaces such as FSL and SPM for motion correction, slice timing correction, spatial normalization, and harmonization. Let *X*_*i*_ *∈* ℝ^*T×V*^ represent the raw fMRI signal for subject *i*, where *T* is the number of time points and *V* is the number of brain regions. The transformation : 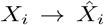 produces a spatially and temporally standardized signal 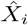. Motion correction is applied by computing the framewise displacement (FD) at each time point, defined as 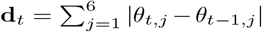, where *θ*_*t,j*_ are the motion parameters. Subjects exceeding an FD threshold of 0.5 mm are excluded. Slice timing correction follows, performed using the FSL SliceTimer interface. Spatial normalization is then applied, transforming *X*_*i*_ to MNI152 space using an affine matrix **A** and a non-linear deformation field **Φ**, such that 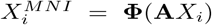. ComBat harmonization adjusts voxelwise intensities *X*_*i,v*_ with 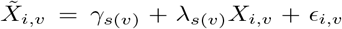, where *s*(*v*) represents the scanner site. Finally, temporal bandpass filtering (0.01–0.1 Hz) is performed to suppress physiological noise while preserving neuronal activity. The resulting time series 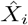 is denoised, aligned, and harmonized for mFC analysis.

#### 2) Modular Brain Mapping

We segment the brain into functionally distinct subnetworks to improve the biological interpretability of functional connectivity. In contrast to wholebrain network construction, modular mapping excludes spurious connections, ensuring that only functionally coherent regions contribute to the analysis. We use the Yeo2011^1^ functional atlas, which defines seven large-scale networks, and the Brainnetome Atlas^2^, which provides fine-grained anatomical parcellation with 246 regions of interest (ROIs). The mapping process involves spatial registration, probabilistic ROI assignment, and partial volume correction to improve spatial accuracy. This modular framework enables functional connectivity analysis sensitive to localized disruptions, enhancing the detection of disorder-specific alterations. Given a pre-processed fMRI dataset 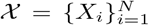, where *X*_*i*_ *∈* ℝ^*T ×V*^ represents the fMRI time series for subject *i*, we define a mapping function *ℳ* that assigns each voxel *v* to a modular network *m* using the spatial probability distributions of the Yeo2011 (*𝒜*_*F*_) and Brainnetome (*𝒜*_*A*_) atlases. The process includes spatial transformations, probabilistic assignments, and partial volume corrections. First, we register (*𝒜*_*A*_ to (*𝒜*_*F*_ resolution via B-spline interpolation, affine transformation **A**, and nonlinear deformation field **Φ**:

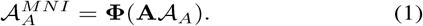

Next, we assign each voxel *v* to a modular network *m* using a probabilistic mapping function *𝒫* (*v, m*):

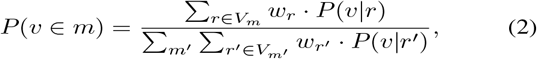

where *P* (*v* | *r*) represents voxel-ROI association, and *w*_*r*_ is a spatial weighting factor based on the Yeo2011 atlas. To correct for boundary voxel misassignment, we apply partial volume correction (PVC) using the weighted overlap function *O*(*v, m*):

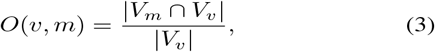

where *V*_*m*_ is the set of voxels in module *m*, and *V*_*v*_ is the voxel neighborhood. Voxels with *O*(*v, m*) *< τ* (*τ* = 0.5) are reclassified based on their nearest high-confidence neighbor. To validate modular assignments, we compute the Dice coefficient *D*(*m*):

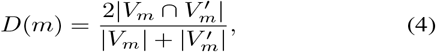

where 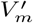 represents the module’s reference segmentation. Assignments with *D*(*m*) *>* 0.7 are retained, ensuring high spatial alignment across subjects.

#### 3) Modular Functional Connectivity Estimation

We estimate mFC by identifying statistically significant interactions within functionally segmented brain subnetworks. Unlike whole-brain connectivity approaches, this method isolates functionally relevant interactions while minimising noise from global fluctuations. We extract representative regional time series, compute direct functional interactions using a sparse graphical model, and apply statistical thresholding to retain only significant connections. The resulting modular connectivity graphs serve as input for subsequent graph-based learning models. Given a modular brain-mapped fMRI dataset 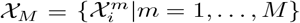, where 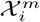 represents the time series of module *m* for subject *i*, we estimate modular connectivity 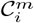 for each network. First, we extract representative time series 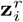 for each region of interest (ROI) *r* using principal component analysis (PCA):

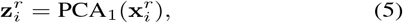

where 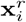 denotes the voxel-level time series. We then apply temporal bandpass filtering (0.01 *≤ f ≤* 0.1 Hz) to suppress non-neuronal fluctuations. Next, we compute the partial correlation matrix 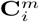, representing direct statistical dependencies between ROIs while controlling for indirect effects. We estimate 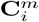 using Graphical Lasso (GLasso), a sparse inverse covariance method with *L*_1_-regularization:

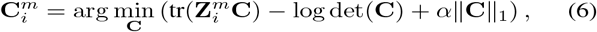

where 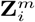 is the sample covariance matrix, *α* (set to 0.01) controls sparsity, and *∥***C***∥*_1_ enforces regularization. This approach retains statistically meaningful interactions while suppressing weak or indirect connections. To ensure robustness, we apply permutation-based significance testing. We generate a null distribution by shuffling time series segments 1,000 times and computing a corresponding connectivity matrix 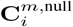. Connectivity edges are thresholded using the 95th percentile of this distribution:

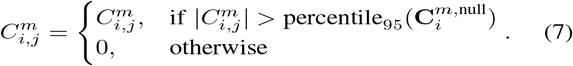

The final mFC representation consists of weighted adjacency matrices 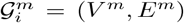, where nodes *V*^*m*^ correspond to ROIs within module *m*, and edges *E*^*m*^ encode significant functional connections. We further extract graph-theoretic features, including degree centrality 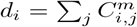 and betweenness centrality *b*_*i*_, to enhance network characterization.

### C. Modular Brain Graph Transformer

We introduce the modular brain graph transformer, a novel architecture for brain disorder classification using mFC patterns. This model integrates self-attention and cross-attention mechanisms to capture localized features within individual modules and interactions across modules.

#### 1) Input Embeddings

We encode modular brain networks into structured graph embeddings to enhance the learning of mFC patterns. Each brain network is represented as a set of modular graphs, where nodes correspond to regions of interest (ROIs) and edges encode functional connectivity. Unlike vectorized fMRI features, these embeddings preserve both functional and topological properties. Given modular graphs 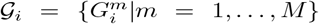, where each module 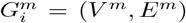, we compute node and edge embeddings. For each node *v*, the initial feature representation is:

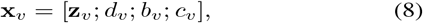

where **z**_*v*_ is the principal component of the node’s fMRI time series, *d*_*v*_ is degree centrality, *b*_*v*_ is betweenness centrality. We normalise all node features using z-score transformation. To encode edge representations, we integrate connectivity strength with self-attention mechanisms. The attention bias for edge *e*_*ij*_ is:

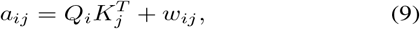

where *Q*_*i*_ and *K*_*j*_ are learnable query and key embeddings, and *w*_*ij*_ is the original edge weight. This ensures that node interactions are conditioned on topological connectivity and learned dependencies.

#### 2) Positional Encoding

We incorporate positional encoding to maintain anatomical consistency across subjects and ensure node embeddings retain spatial information relevant to brain network organisation. Unlike sequential positional encodings in natural language processing, brain networks require a spatially informed representation reflecting anatomical structure. We use Laplacian positional encoding (LPE) to capture spatial dependencies and introduce learnable spatial embeddings initialized from MNI coordinates to refine spatial representation during training. We define a spatial embedding function *𝒮* : *V*^*m*^*→* ℝ^*d*^ that assigns each node a position-aware vector. First, we compute Laplacian positional encoding using the graph Laplacian **L** = **D** *−***A**, where **D** is the degree matrix and **A** is the adjacency matrix. The eigen-decomposition of **L** is given by **LU** = **UΛ**, where **U** contains eigenvectors, and **Λ** is the diagonal matrix of eigenvalues. We select the first *k* = 8 eigenvectors corresponding to the smallest nonzero eigenvalues to define LPE as **p**_*v*_ = **U**_*k*_(*v*). To enhance spatial representation, we introduce learnable spatial embeddings initialized with MNI coordinates **x**_*v*_ = (*x*_*v*_, *y*_*v*_, *z*_*v*_). The learnable spatial embedding is:

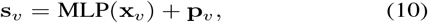

where the multi-layer perceptron (MLP) transforms MNI coordinates into a high-dimensional space, refining spatial encoding. The final position-aware node embedding is:

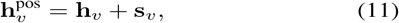

where **h**_*v*_ is the original node embedding. This formulation ensures that graph representations integrate both functional and spatial constraints.

#### 3) Modular Encoders

We employ modular encoders to extract localized connectivity patterns within each brain subnetwork independently. Unlike whole-brain graph learning, which aggregates features indiscriminately, this approach preserves intrinsic connectivity structures while eliminating irrelevant global interactions. By leveraging Performer (FAVOR+) self-attention [44], we reduce computational complexity from *O*(*n*^2^) to *O*(*n*), enabling scalable modular graph learning. Each node *v* in module *m* has an initial embedding 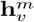, integrating functional, topological, and positional features. We compute self-attention scores within modules using:

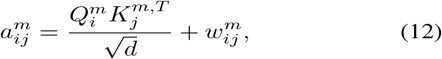

where 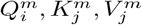 are query, key, and value projections of node embeddings 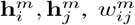 is the edge weight, and *d* is the feature dimension. We mask weak connections by setting attention scores to zero for edges with *p <* 0.01, reducing noise. The updated node representation is:

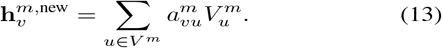

To enhance disorder-specific representation learning, we introduce module-specific parameters for functionally critical subnetworks, such as the default mode network (DMN), salience network (SN), and limbic network (LN). For these, we learn independent transformation matrices 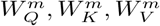, while sharing parameters across less disorder-relevant modules. The final output is a set of modular network embeddings: 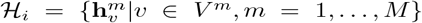 capturing localized functional connectivity properties within each module.

#### 4) Cross-Modular Encoders

We model inter-network interactions using cross-attention mechanisms to capture dependencies between functionally distinct brain subnetworks. Modular encoders extract localised connectivity patterns, while cross-modular encoders learn higher-order interactions, crucial for understanding cognitive integration and disease pathology. Neurological disorders often disrupt cross-network communication rather than isolated subnetworks, necessitating an approach that preserves both local and global connectivity features. To enable structured information flow across subnetworks, we employ a cross-attention mechanism while maintaining network-specific integrity. We define the cross-attention score between node *i* in module *m* and node *j* in module *n* as:

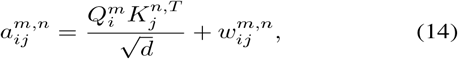

where 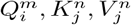 are query, key, and value projections,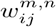 represents inter-module connectivity strength, and *d* is the feature dimensionality. To prevent feature dilution, we introduce a global classification token ([CLS]) aggregating cross-modular dependencies:

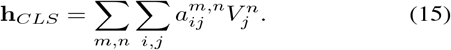

This token provides a global representation of inter-network communication, refined through hierarchical pooling. To maintain modular integrity and enhance feature propagation, we incorporate residual connections:

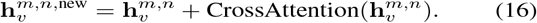

These connections preserve modular structure while enriching representations with inter-network dependencies. The final output, 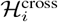, encapsulates intra- and inter-network relationships, improving sensitivity to neurodegenerative and neurodevelopmental disruptions. By integrating global attention pooling, our approach enhances interpretability of large-scale brain network interactions, facilitating precise characterization of disease-specific connectivity signatures.

#### 5) Adaptive Fusion and Classification

We integrate cross-module interaction features into a hierarchical attention mechanism to classify brain disorders. Unlike traditional graph-based models that treat brain networks as a single entity, our approach fuses localised and cross-modular connectivity patterns, improving classification accuracy. This method preserves local connectivity disruptions while capturing global alterations in inter-network communication. Hierarchical attention pooling dynamically weighs subnetwork contributions, enhancing disorder-relevant feature selection. Given cross-modular interaction features, 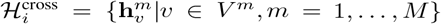, we compute attention weights *α*_*m*_ for each module:

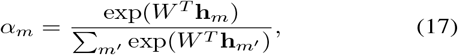

where *W* is a learnable weight vector and **h**_*m*_ is the aggregated module embedding. The global representation is computed as:

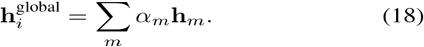

This mechanism prioritises informative subnetworks while suppressing redundant features. The global embedding is passed through a multi-layer perceptron (MLP) classifier:

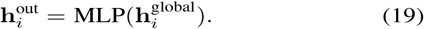

Each layer applies GeLU activation to enhance gradient flow and prevent saturation. The final classification output *ŷ*_*i*_ is obtained using softmax:

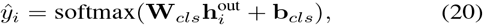

where **W**_*cls*_ and **b**_*cls*_ are learnable classification parameters. To mitigate class imbalances, we employ Focal Loss:

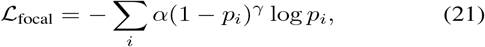

where *p*_*i*_ is the predicted probability for the correct class, *γ* = 2.0 controls loss focusing, and *α* balances class weights. This improves generalisation by emphasising underrepresented classes in imbalanced medical datasets. The pseudocode of mBrainGT is given in Algorithm 1.

##### Algorithm 1 mBrainGT Algorithm

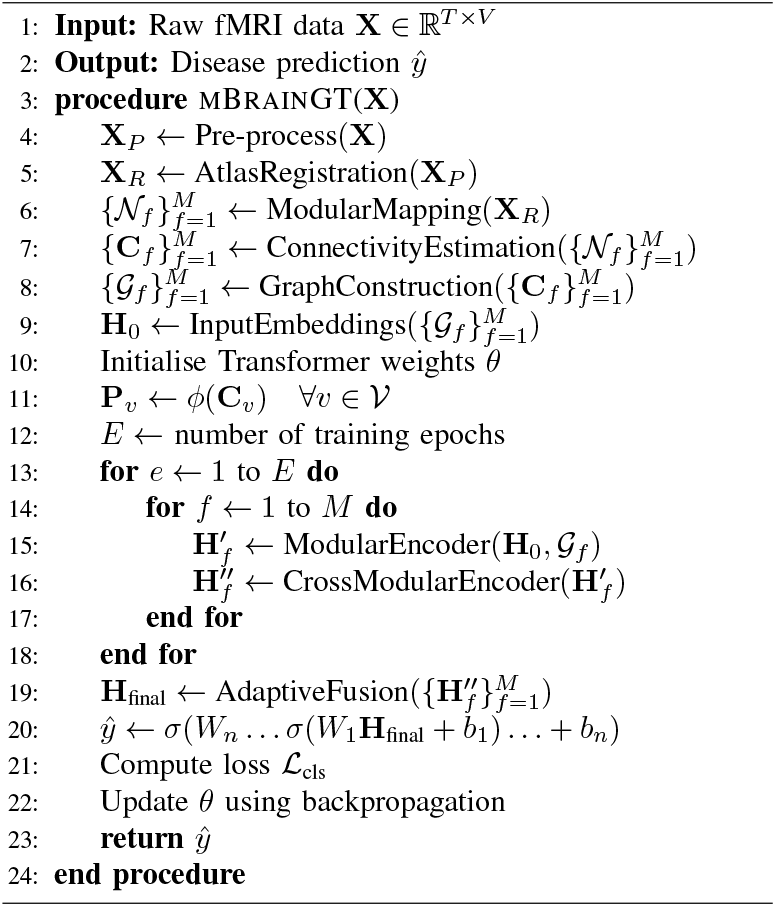

## IV. Experimental Results

In this section, we evaluate the effectiveness of our proposed mBrainGT model through comprehensive experiments. Our evaluation focuses on addressing the following research questions:

RQ1: How does mBrainGT perform in diagnosing brain disorders across ADNI, PPMI, and ABIDE datasets?

RQ2: What improvements in classification metrics does mBrainGT offer over traditional whole-brain connectivity methods and other baselines?

RQ3: What is the impact of different components in mBrainGT on the overall performance and diagnostic accuracy?

RQ4: What are the optimal parameters for the mBrainGT model to achieve the highest diagnostic accuracy?

RQ5: How interpretable are the modular brain network patterns identified by mBrainGT in relation to brain disorders?

### A. Experimental Settings

#### 1) Datasets

1. *ADNI Dataset*, namely Alzheimer’s Disease Neuroimaging Initiative (ADNI)^3^ dataset provides fMRI data for studying Alzheimer’s disease (AD) progression. Researchers can access the dataset upon application approval for neuroimaging and biomarker analysis. The primary objective is to identify biomarkers for AD detection and monitoring. Subjects are categorised as cognitively normal, mild cognitive impairment, or AD. A subset of 426 participants includes 199 females (46.7%) and 146 AD patients (34.2%), with ages ranging from 50 to 100 years. The selection process ensures the inclusion of subjects with confirmed diagnoses, maintaining data integrity for analysis.
2. *PPMI Dataset*, namely The Parkinson’s Progression Markers Initiative (PPMI)^4^ dataset, available on the PPMI website, contains fMRI data to study Parkinson’s Disease (PD) progression. Researchers meeting specific criteria can access this dataset to explore PD biomarkers. The dataset aims to identify diagnostic and progression markers for PD, distinguishing between healthy controls and PD patients. The study cohort includes fMRI data from 823 individuals, with 230 (70.9%) diagnosed with PD, 130 (40.1%) female, and ages 40 to 85, representing a cross-section of the PD-affected population.
3. *ABIDE Dataset*, namely The Autism Brain Imaging Data Exchange (ABIDE)^5^ dataset provides fMRI data for Autism Spectrum Disorder (ASD) research. This dataset, sourced from multiple international locations, aims to elucidate ASD neural underpinnings and identify diagnostic biomarkers. Participants are categorised into ASD or control groups. It includes fMRI data from 1118 subjects aged 8 to 40 years, engaged in social cognition tasks. Of these, 537 (48%) are diagnosed with ASD and 161 (14.5%) are female. A detailed summary of the datasets is presented in Table I.

**TABLE I.** The Statistices of Datasets.

| Dataset | Subjects | Age Range | Male/Female | Labels |
| --- | --- | --- | --- | --- |
| ADNI | 426 | 50-100 | 227/199 | CN (280), AD (146) |
| PPMI | 324 | 40-85 | 194/130 | Control (94), PD (230) |
| ABIDE | 1118 | 8-40 | 957/161 | Control (581), ASD (537) |

#### 2) Baselines

##### a) Conventional Machine Learning Models

Conventional machine learning models establish baseline performance for feature-driven approaches. We employ Support Vector Machine (SVM), Random Forest (RF), and Multi-Layer Perceptron (MLP) using handcrafted graph-theoretic features derived from whole-brain functional connectivity matrices. These features encode network topology, nodal centrality, and connectivity strength, enabling neuropsychiatric classification. Brain networks are constructed via the BrainGB preprocessing pipeline, and extracted features serve as model inputs. We train and optimise models using scikit-learn Python library, applying grid search and cross-validation for hyperparameter tuning to ensure robust generalization across datasets.

##### c) Conventional Graph Learning Models

Conventional graph learning models extend traditional models by directly processing brain network structures. We evaluate three architectures: Graph Isomorphism Network (GIN), Graph Attention Network (GAT), and Graphormer. These models perform end-to-end representation learning on functional connectivity-derived brain graphs without explicit feature selection. We generate brain networks using the BrainGB pipeline and implement models with PyTorch Geometric to ensure consistent graph processing. Unlike conventional approaches, GNNs dynamically learn node embeddings and graph representations, enabling flexible adaptation to complex neuroimaging data distributions.

##### c) Brain Network Models

Brain network models are graph-based architectures tailored for neuroimaging applications, integrating domain-specific constraints to improve interpretability and predictive performance. We evaluate BrainGNN, BrainGB, and KDGCN [34] as baselines. BrainGNN employs ROI-aware graph convolutional layers and ROI-selection pooling for adaptive feature extraction from functionally relevant regions. BrainGB offers a benchmark framework supporting diverse GNN architectures for systematic evaluation. KDGCN applies a knowledge distillation-guided learning paradigm with a teacher-student framework to enhance classification accuracy while preserving neurobiological insights. All models are trained on functional connectivity graphs using official configurations, pre-processing protocols, and original hyperparameters.

##### d) Brain Graph Transformers

Transformer-based models leverage self-attention mechanisms to capture long-range dependencies and hierarchical interactions in brain networks. We benchmark mBrainGT against BRAINNETTF, ALTER [40], and Medical Transformer [41]. BRAINNETTF employs a transformer encoder with orthonormal clustering readout for structured representation learning. ALTER integrates a biased random-walk strategy to model multi-scale neural interactions. Medical Transformer processes 3D brain images as 2D sequences using multi-view spatial encoding and self-supervised pre-training. All models are trained on functional connectivity networks with official hyperparameters and optimisation strategies, ensuring methodological consistency.

#### 3) Evaluation Metrics

The efficacy of the mBrainGT framework was evaluated using accuracy, F1-score, and AUC. Accuracy reflects overall performance, while F1-score balances precision and recall, crucial for imbalanced datasets. AUC assesses performance across thresholds, ensuring robustness in brain disease diagnosis. A paired t-test (*α* = 0.05) verified that performance gains were statistically significant and not due to random variation.

#### 4) Implementation Details

The mBrainGT framework, developed in PyTorch, comprises two principal components: the modular brain network construction module and the modular brain graph transformers module. The former module processes fMRI data through atlas registration, ROI extraction and assignment to functional networks, functional connectivity estimation, and construction of modular brain network datasets. The latter module embeds these networks, implements positional encoding, utilises network-specific and cross-network encoders, and adaptively fuses the output for classification purposes. Data were partitioned into training, validation, and testing subsets with proportions of 70%, 15%, and 15%, respectively. Training was performed using the Adam optimizer, configured with a learning rate of 0.001, a batch size of 32, and stopping criteria set at 100 epochs. Hyperparameter tuning was performed using a grid search approach. All computational experiments were carried out on a workstation equipped with an NVIDIA 4090 RTX Ti 48 GB GPU.

#### 5) Computational Complexity

We analyse the time and space complexity of mBrainGT by examining key operations, including matrix computations, partial correlation estimation, graph processing, and self-attention mechanisms. Each operation’s complexity is derived using Big-O notation. The most computationally intensive steps involve mFC estimation and the graph transformer. Partial correlation and self-attention mechanisms scale quadratically with the number of regions of interest (ROIs), denoted as *P*, resulting in an overall time complexity of *O*(*P*^2^). The highest memory consumption stems from storing brain networks, adjacency matrices, and intermediate graph embeddings, each scaling as *O*(*P*^2^). Thus, both time and space complexities follow a quadratic relationship with *P*, making the model computationally demanding but scalable in terms of the number of modules and subjects processed.

### B. Performance of mBrainGT

We assess the performance of mBrainGT in addressing RQ1 by evaluating classification accuracy, F1-score, and AUC (Table II). The model demonstrates high accuracy across datasets, peaking in PPMI (85.42 ± 2.89%), followed by ABIDE (82.74 ± 4.53%) and ADNI (74.92 ± 2.34%). These results confirm the robustness of mFC in differentiating neurological conditions. However, AUC trends reveal key insights. The highest AUC occurs in ABIDE (85.84 ± 8.12%), indicating that mFC effectively captures localized disruptions in ASD. In contrast, PPMI achieves the highest accuracy but a lower AUC (68.17 ± 5.22%), suggesting suboptimal confidence in distinguishing Parkinson’s disease (PD) from controls. The lower F1-scores across datasets (65.67%–70.68%) suggest classification bias or class imbalance. Particularly in PPMI, this may reflect the complexity of class separability. Performance variability in ABIDE further highlights high inter-subject heterogeneity in ASD phenotypes. These findings validate mBrainGT’s diagnostic potential while identifying areas for improvement in feature sensitivity and disorder-specific generalization.

**TABLE II.** Comparative Performance Analysis (%). mBrainGT demonstrates statistically significant improvements over baseline models, confirmed by t-tests (p-value *<* 0.05).

| Type | Method | ADNI |  |  | PPMI |  |  | ABIDE |  |  |
| --- | --- | --- | --- | --- | --- | --- | --- | --- | --- | --- |
|  |  | Accuracy | F1-score | AUC | Accuracy | F1-score | AUC | Accuracy | F1-score | AUC |
| Machine Learning Models | SVM | 60.34±0.17 | 42.44±0.17 | 62.58±4.80 | 74.09±0.24 | 44.21±0.08 | 49.21±1.48 | 59.4±2.3 | 46.7±24.0 | 62.4±3.5 |
|  | Random Forest | 60.51±2.85 | 45.36±2.86 | 63.38±3.50 | 77.02±1.97 | 49.45±3.51 | 58.51±0.11 | 51.2±5.4 | 43.9±34.2 | 55.3±1.9 |
|  | MLP | 61.90±1.22 | 43.14±0.57 | 65.74±2.37 | 77.99±2.78 | 52.96±4.52 | 59.65±6.80 | 60.7±12.6 | 51.1±40.9 | 66.7±3.8 |
| Graph Learning Models | GIN | 65.85±6.42 | 52.29±5.15 | 69.36±5.09 | 78.41±1.62 | 54.46±3.08 | 63.24±3.77 | 63.6±1.9 | 53.7±8.3 | 69.7±3.3 |
|  | GAT | 64.63±2.35 | 57.12±1.81 | 65.32±1.42 | 79.34±2.21 | 55.98±3.08 | 64.15±2.67 | 67.8±2.7 | 58.8±9.7 | 74.9±2.4 |
|  | Graphormer | 67.62±4.85 | 58.93±4.01 | 70.17±3.23 | 78.42±3.53 | 56.55±6.48 | 63.07±7.75 | 71.2±1.2 | 60.4±11.4 | 75.4±1.2 |
| Brain Network Models | BrainGNN | 64.93±4.90 | 55.94±5.42 | 67.94±5.42 | 77.30±1.52 | 50.96±4.20 | 62.93±4.89 | 72.4±2.3 | 56.7±24.0 | 76.4±3.5 |
|  | BrainGB | 67.09±2.77 | 64.09±2.77 | 73.36±4.26 | 80.52±6.23 | 57.45±6.28 | 63.09±3.25 | 76.3±6.3 | 62.7±34.0 | 78.4±3.7 |
|  | KDGCN | 71.97±0.13 | 63.92±0.06 | 72.00±0.20 | 78.91±3.61 | 56.86±2.68 | 63.14±5.37 | 78.9±8.2 | 61.4±15.0 | 79.5±6.3 |
| Brain Graph Transformers | BRAINNETTF | 70.46±0.82 | 65.79±0.83 | 74.18±1.16 | 81.39±0.52 | 58.68±3.12 | 64.29±2.11 | 77.4±1.7 | 62.4±7.8 | 78.5±5.4 |
|  | ALTER | 72.92±1.29 | 67.45±1.28 | 76.16±1.41 | 82.29±2.58 | 62.59±2.1 | 64.35±4.71 | 80.4±5.3 | 64.4±7.8 | 82.5±5.4 |
|  | Medical Transformer | 71.42±0.11 | 66.30±2.08 | 74.33±0.78 | **86.43±1.92** | 63.43±4.6 | 65.23±2.79 | 79.2±4.9 | 65.6±16.0 | 83.2±4.5 |
| Ours | mBrainGT | **74.92±2.34** | **70.68±1.86** | **78.87±3.21** | 85.42±2.89 | **65.67±1.57** | **68.17±5.22** | **82.74±4.53** | **67.93±2.90** | **85.84±8.12** |

### C. Comparison with Baseline Methods

To address RQ2, we assess the performance of mBrainGT against established baseline methods. Table II presents the classification results across three datasets (ADNI, PPMI, and ABIDE), quantifying the improvements mBrainGT offers in accuracy, F1-score, and AUC.

#### 1) Conventional Machine Learning Models

Across all datasets, mBrainGT surpasses conventional machine learning models in classification performance. The best-performing baseline, MLP, achieves accuracies of 61.90% (ADNI), 77.99% (PPMI), and 60.7% (ABIDE). In contrast, mBrainGT improves accuracy by 22.04% (ADNI), 7.43% (PPMI), and 22.04% (ABIDE). It also yields higher F1-scores and AUC values, demonstrating superior identification of disorder-specific connectivity patterns. Conventional models rely on handcrafted features, which introduce information loss and fail to capture topological complexities in functional brain networks.

#### 2) Comparison with Graph Learning Models

Graph learning models, including GIN, GAT, and Graphormer, represent functional connectivity by modeling the brain as a graph, where nodes correspond to regions of interest (ROIs) and edges encode functional interactions. These models preserve topological properties and enable node-wise message passing, making them effective for connectivity-based classification. Among them, Graphormer achieves the highest accuracy: 67.62% (ADNI), 79.34% (PPMI), and 71.2% (ABIDE). However, mBrainGT outperforms Graphormer by 11.54% (ADNI), 6.99% (PPMI), and 11.54% (ABIDE). This advantage extends to F1-score and AUC, highlighting the benefits of mFC. Traditional graph learning models rely on whole-brain connectivity graphs, making them vulnerable to noisy edges and redundant connections. mBrainGT mitigates these issues by constructing modular graph representations, capturing both intra-network and inter-network disruptions.

#### 3) Brain Network Models

Brain network models, including BrainGNN, BrainGB, and KDGCN, incorporate neuroscientific priors by emphasizing disorder-relevant ROIs and connectivity structures. These models improve interpretability and clinical relevance by integrating biological constraints. Among them, KDGCN achieves the highest accuracy: 71.97% (ADNI), 78.91% (PPMI), and 78.9% (ABIDE). However, mBrainGT surpasses KDGCN by 3.84% (ADNI), 5.71% (PPMI), and 3.84% (ABIDE). The improvements in F1score and AUC indicate superior classification robustness and generalizability. Traditional brain network models prioritize specific ROIs but often discard global contextual information, limiting adaptability to heterogeneous disorder manifestations. mBrainGT addresses this limitation through modular representation learning, preserving local specificity while maintaining global dependencies. Its adaptive fusion of intra- and inter-modular interactions enhances functional disruption modeling, improving diagnostic reliability.

#### 4) Brain Graph Transformers

Transformer-based models, including BRAINNETTF, ALTER, and Medical Transformer, utilise self-attention to capture long-range dependencies in brain networks. These models dynamically weight functional connections based on disorder-relevant patterns, enhancing classification performance. Among them, ALTER achieves the highest accuracy: 72.92% (ADNI), 82.29% (PPMI), and 80.4% (ABIDE). However, mBrainGT surpasses ALTER by 2.34% (ADNI), 1.01% (PPMI), and 2.34% (ABIDE). The gains in F1-score and AUC further highlight its advantage in modular connectivity-aware attention. While transformers effectively model global dependencies, they lack disorder-specific constraints and treat all ROIs uniformly, amplifying non-informative connections. mBrainGT addresses this by integrating modular priors, selectively focusing on clinically relevant subnetworks. Comparative analysis confirms mBrainGT consistently outperforms baseline models across ADNI, PPMI, and ABIDE. Its ability to capture disorder-relevant modular connectivity patterns reduces noise and enhances interpretability. By combining self-attention with adaptive fusion, mBrainGT achieves state-of-the-art diagnostic accuracy.

### D. Ablation Study

We conduct an ablation study to evaluate the contribution of individual components to the mBrainGT framework’s diagnostic accuracy (RQ3). The baseline model includes all components: mFC, modular encoders, cross-modular encoders, positional encoding, and adaptive fusion. We systematically remove each component to assess its impact on performance. Ablation scenarios include the exclusion of modular brain networks, modular encoders, cross-modular encoders, positional encoding, and adaptive fusion. Additionally, we evaluate a core model retaining only the basic graph transformer architecture. All experiments use the ADNI, PPMI, and ABIDE datasets with consistent training, validation, and test splits. We assess model performance using accuracy, F1-score, and AUC, ensuring a comprehensive evaluation of its diagnostic capability.

Our results demonstrate the impact of each ablation scenario on diagnostic accuracy (Table III). On the ADNI dataset, removing modular brain networks reduces accuracy by 4.14% (72.76% to 68.62%), highlighting their role in capturing functional interactions. Eliminating modular encoders leads to the most significant drop (7.28%), underscoring their necessity for analyzing brain subnetworks. Removing cross-modular encoders results in a smaller 2.60% decrease, suggesting their supportive but non-critical role. Excluding adaptive fusion reduces accuracy by 1.46%, indicating its minor contribution. The full ablation causes a 14.55% decline, confirming the model’s dependence on all components. On the PPMI dataset, removing modular networks lowers accuracy by 4.54% (78.89% to 74.35%), while modular encoder removal results in the largest 7.92% drop. Cross-modular encoders and adaptive fusion contribute marginally, reducing accuracy by 1.76% and 0.79%, respectively. The full ablation decreases accuracy by 15.78%, reinforcing the framework’s necessity. On ABIDE, modular network removal decreases accuracy by 3.26% (76.47% to 73.21%), and modular encoder removal causes a 7.66% drop. Excluding cross-modular encoders and adaptive fusion reduces accuracy by 2.50% and 2.29%, respectively. The largest decline (15.29%) occurs with full ablation, confirming the framework’s critical role in ASD diagnosis.

**TABLE III.** Performance Impact of Component Ablation on mBrainGT Framework: A Comparative Analysis Across adni, ppmi, and ABIDE Datasets.

| Component Ablated | ADNI |  |  | PPMI |  |  | ABIDE |  |  |
| --- | --- | --- | --- | --- | --- | --- | --- | --- | --- |
|  | Accuracy | F1-score | AUC | Accuracy | F1-score | AUC | Accuracy | F1-score | AUC |
| None (Full mBrainGT) | 74.92 | 70.68 | 78.87 | 85.42 | 65.67 | 68.17 | 82.74 | 67.93 | 85.84 |
| w/o modular brain networks | 68.62 | 63.43 | 73.17 | 74.35 | 62.66 | 64.63 | 73.21 | 65.89 | 75.31 |
| w/o modular encoders | 65.48 | 59.99 | 68.71 | 70.97 | 59.23 | 61.31 | 68.81 | 63.14 | 70.83 |
| w/o cross-modular encoders | 70.16 | 65.05 | 74.08 | 77.13 | 63.19 | 66.21 | 73.97 | 68.25 | 75.71 |
| w/o adaptive fusion | 71.30 | 66.24 | 73.16 | 78.10 | 62.85 | 66.78 | 74.18 | 69.35 | 77.15 |
| All Components | 58.21 | 54.04 | 60.33 | 63.11 | 53.31 | 53.83 | 61.18 | 56.74 | 62.20 |

Our ablation study highlights the critical role of modular encoders and modular brain networks in diagnostic accuracy. Modular encoders, which analyse brain subnetworks, have the most significant impact, with their removal causing the largest accuracy drop, particularly in the PPMI dataset. Modular brain networks also improve performance by capturing subnetwork interactions. Cross-modular encoders and adaptive fusion enhance accuracy but contribute less critically, as their removal results in smaller declines. The largest performance drop occurs when all components are removed, underscoring the framework’s holistic design. Each component plays a distinct role in capturing complex brain network disruptions essential for accurate diagnosis.

### E. Parameter Analysis

To optimise mBrainGT for brain disorder diagnosis (RQ4), we conducted a parameter analysis focusing on the number of functional networks (*n*) and encoder layers (*l*). We varied *n* from 1 to 17, covering predefined Yeo2011 functional networks, and adjust *l* from 1 to 20 to determine optimal encoder depth. Experiments utilised ADNI, PPMI, and ABIDE datasets, with baseline settings derived from preliminary trials. We systematically modified *n* and *l* while tracking accuracy and F1-score. Performance curves visualize the impact of parameter variations, offering insights into model efficacy.

Our findings reveal a non-linear relationship between functional networks (*n*) and model performance, with improvements plateauing beyond an optimal range. The results are shown in Fig. 4, and Fig. 3. Increasing encoder layers (*l*) yields diminishing returns, indicating a trade-off between complexity and computational efficiency. Optimal parameters are *n* = 11, *l* = 2 (ADNI), *n* = 9, *l* = 3 (PPMI), and *n* = 14, *l* = 2 (ABIDE), selected based on accuracy and F1-score. The optimal *n* and *l* values vary across datasets and brain disorders, highlighting the necessity of dataset-specific tuning. While these results provide locally optimal configurations, further parameter exploration is needed to enhance diagnostic performance.

**Fig. 3.**
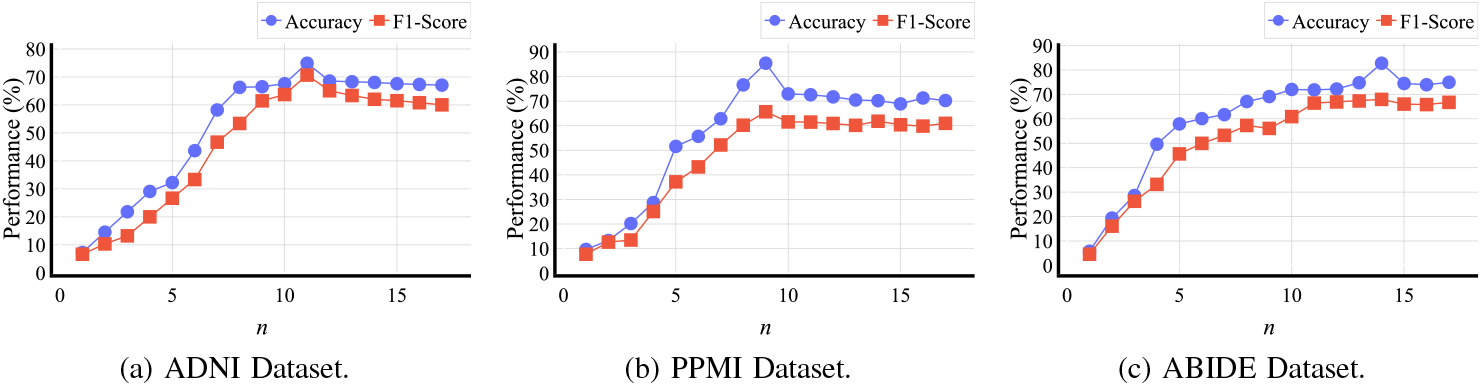
The accuracy and F1-score of mBrainGT w.r.t. different *n* values.

**Fig. 4.**
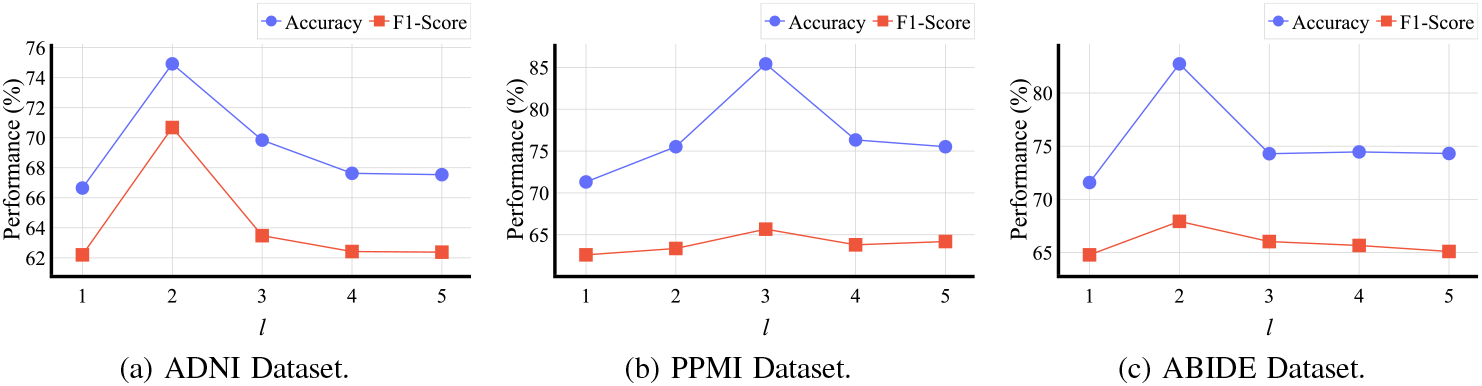
The accuracy and F1-score of mBrainGT w.r.t. different *l* values.

### F. Interpretability Analysis

To evaluate the interpretability of mBrainGT (RQ5), we conduct a SHapley Additive exPlanations (SHAP) analysis to assess the contribution of functional networks in diagnosing Alzheimer’s Disease (AD), Parkinson’s Disease (PD), and Autism Spectrum Disorder (ASD). We apply SHAP to the ADNI, PPMI, and ABIDE datasets, treating brain modules as features and deriving their importance scores. This framework quantifies each network’s contribution to model predictions, providing insights into disorder-specific biomarkers. By computing SHAP values for each dataset, we identify the most critical modular brain networks for accurate diagnosis.

The SHAP summary plots (Fig. 5) illustrate feature importance for each dataset and task. For AD detection, the Default Mode Network (DMN) exhibits the highest SHAP values, confirming its critical role in Alzheimer’s pathology. DMN disruptions, widely linked to cognitive deficits in memory and self-referential thinking, align with prior studies. For PD detection, the Somatomotor Network (SMN) shows the highest SHAP values in the PPMI dataset, supporting evidence of motor dysfunctions as a primary Parkinson’s hallmark. Given its role in sensorimotor control, SMN connectivity disruptions are expected in PD. For ASD detection, the Visual Network (VN) has the greatest impact, as reflected in the ABIDE dataset, consistent with literature on sensory and visual processing abnormalities in ASD. The interpretability analysis confirms that mBrainGT identifies disorder-specific brain networks, enhancing clinical relevance [21], [45], [46]. Its modular approach isolates network-specific disruptions, offering a more refined understanding than whole-brain connectivity methods, which often obscure localized alterations [19].

**Fig. 5.**
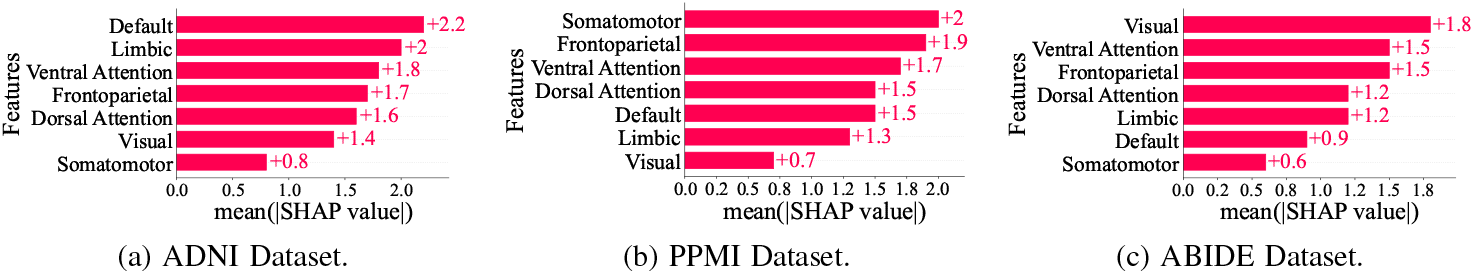
The accuracy and F1-score of mBrainGT w.r.t. different *n* values.

### G. Discussion

The results of mBrainGT validate the hypothesis that mFC analysis improves diagnostic accuracy by reducing noise from non-informative connections and identifying localized pathological disruptions critical for disorder-specific signatures. The model demonstrates high classification accuracy across three major neuroimaging datasets (ADNI, PPMI, ABIDE), indicating its robustness in distinguishing neurological conditions. Notably, the model achieves an AUC of 85.84% for ASD classification, highlighting the efficacy of modular connectivity in capturing functional disruptions specific to ASD. In contrast, the lower AUC in PD classification (68.17%) suggests that subtle neurodegenerative patterns in Parkinson’s disease challenge modular analysis, necessitating advanced feature extraction methods. The ablation study confirms the importance of self-attention and cross-attention mechanisms, as performance significantly declines when these components are removed. These findings demonstrate the advantage of brain modular network representations over whole-brain connectivity approaches, addressing limitations of traditional graph learning models prone to noise and redundancy.

The findings of mBrainGT have significant implications for computational neuroscience, clinical diagnostics, and personalized medicine. mBrainGT shifts from whole-brain connectivity models to a modular framework, emphasizing functional disruptions within disease-relevant brain networks. This modular approach enhances understanding of neurological disorders at the subnetwork level, providing a biologically interpretable method for brain network analysis. By incorporating self-attention and cross-attention mechanisms, mBrainGT captures both local dysfunction and system-level alterations in neurodegenerative and neurodevelopmental disorders. This is particularly relevant for Alzheimer’s disease and ASD, where localized network changes contribute to cognitive decline or developmental abnormalities. Clinically, mBrainGT’s enhanced accuracy suggests its potential for early diagnosis, prognosis monitoring, and treatment evaluation, especially when integrated with multimodal neuroimaging tools. However, challenges remain in scalability, dataset generalizability, and computational complexity. Future work should focus on more efficient architectures and multi-modal data integration to improve generalization, clinical applicability, and interpretability. Addressing these limitations will be key for mBrainGT’s broader clinical utility.

## V. Conclusion

In this study, we address the limitations of whole-brain connectivity methods in diagnosing brain disorders by introducing the mBrainGT framework, which emphasises mFC. This innovative approach focuses on capturing localized disruptions within specific brain subnetworks, enhancing diagnostic accuracy for Alzheimer’s disease (AD), Parkinson’s disease (PD), and autism spectrum disorder (ASD). By leveraging a modular brain graph transformer model, mBrainGT effectively integrates local and global connectivity features through self-attention, cross-attention, and adaptive fusion mechanisms. Experimental results on the ADNI, PPMI, and ABIDE datasets demonstrate superior performance compared to existing methods. Future research will explore dynamic functional connectivity and multiscale analysis approaches to further improve diagnostic precision. Ultimately, this study highlights the value of modular connectivity analysis in providing a more precise and biologically relevant representation of brain network disruptions, offering significant improvements in the diagnosis of brain disorders.

## Data Availability

All data produced in the present study are available upon reasonable request to the authors

## Footnotes

1 https://surfer.nmr.mgh.harvard.edu/fswiki/CorticalParcellation_Yeo2011

2 https://atlas.brainnetome.org/

3 http://adni.loni.usc.edu/data-samples/access-data

4 https://www.ppmi-info.org/access-data-specimens/download-data

5 http://fcon1000.projects.nitrc.org/indi/abide

